# Risk of SARS-CoV-2 testing, PCR-confirmed infections and COVID-19--related hospital admissions in children and young people: birth cohort study

**DOI:** 10.1101/2021.12.17.21267350

**Authors:** Pia Hardelid, Graziella Favarato, Linda Wijlaars, Lynda Fenton, Jim McMenamin, Tom Clemens, Chris Dibben, Ai Milojevic, Alison Macfarlane, Jonathon Taylor, Steven Cunningham, Rachael Wood

## Abstract

**Background:** There have been no population-based studies of SARS-CoV-2 testing, PCR-confirmed infections and COVID-19-related hospital admissions across the full paediatric age range. We examine the epidemiology of SARS-CoV-2 in children and young people (CYP) aged <23 years.

**Methods:** We used a birth cohort of all children born in Scotland since 1997, constructed via linkage between vital statistics, hospital records and SARS-CoV-2 surveillance data. We calculated risks of tests and PCR-confirmed infections per 1000 CYP-years between August and December 2020, and COVID-19-related hospital admissions per 100,000 CYP-years between February and December 2020. We used Poisson and Cox proportional hazards regression models to determine risk factors.

**Results:** Among the 1226855 CYP in the cohort, there were 378402 tests, 19005 PCR confirmed infections and 346 admissions, corresponding to rates of 770.8/1000 (95% confidence interval 768.4-773.3), 179.4 (176.9-182.0) and 29.4/100,000 (26.3-32.8) CYP-years respectively. Infants had the highest COVID-19-related admission rates. Chronic conditions, particularly multiple types of conditions, was strongly associated with COVID-19-related admissions across all ages. The hazard ratio for >1 chronic condition type was 12.2 (7.9-18.82) compared to children with no chronic conditions. 89% of admitted children had no chronic conditions recorded.

**Conclusions:** Infants, and CYP with chronic conditions are at highest risk of admission with COVID-19, however the majority of admitted CYP have no chronic conditions. These results provide evidence to support risk/benefit analyses for paediatric COVID-19 vaccination programmes. Studies examining whether maternal vaccine during pregnancy prevents COVID-19 admissions in infants are urgently needed.

**Funding:** UK Research and Innovation-Medical Research Council

## Background

Children are much less likely to experience hospital admission and mortality related to severe acute respiratory syndrome coronavirus-2 (SARS-CoV-2) infection than adults.^1^ In Europe in 2020, 1.7% of COVID-19-related hospital admissions were in children <19 years of age.^2^

Whilst early reports during the spring and summer of 2020 indicated less than 5% of SARS-CoV-2 positive cases in the United States,^3^ China^4^ and Spain^5^ were in children, the epidemiology has changed over time and place, particularly in association with vaccination policy and access, with children under 10 years of age being the dominant population infected in the latter part of 2021 in the UK..^6^

Over the course of the pandemic our understanding of how SARS-CoV-2 infection affects children has also improved. Children who experience more severe symptoms of SARS-CoV-2 may present with acute infection symptoms such as fever, cough, shortness of breath, nausea/vomiting, or upper respiratory symptoms.^7–9^ Children may also present with an acute inflammatory syndrome, paediatric inflammatory syndrome temporally associated with SARS-CoV-2 (PIMS-TS; also referred to as multisystem inflammatory syndrome related to COVID - MIS-C), several weeks after initial infection.^10–12^ Children aged <2 years old appear to be over-represented among children admitted to hospital with acute symptoms of SARS-CoV-2 infection, whereas children aged 10 years or older account for the largest proportion of admitted PIMS-TS cases.^8 13^

Two preprints indicate that among children admitted to hospital with SARS-CoV-2 or PIMS-TS, those with specific chronic respiratory, neurological, gastrointestinal or cardiovascular conditions, and particularly children with multiple comorbidities, were at increased risk of Paediatric Intensive Care Unit (PICU) admission or death. Infants and teenagers appeared to have higher odds of these severe outcomes compared to children aged 1-4 years old.^14 15^ A lower reported risk of severe disease and, until 2021, relatively lower rates of infection in children have supported a narrative that the benefits and risks (primarily of myocarditis following second dose mRNA vaccines in young men^16 17^) of vaccinations in children are finely balanced.

Most studies of paediatric SARS-CoV-2 infection have been case series of infected or hospitalised children, making calculations of population-based risks of confirmed infections and associated admissions among different groups of children, including children with chronic conditions, impossible. A recent study presented population-based risks of COVID-19-related admission for children with asthma, indicating an increased risk that supports vaccine strategies targeted to this group.^18^ Our aim was to provide population based estimates of risk of SARS-CoV-2 testing, PCR confirmed infections and COVID-19 related admissions in children and young people (CYP) based on age, presence of multiple chronic conditions, and socioeconomic status, that could support vaccination and other policy recommendations across the whole paediatric population.

## Methods

### Data sources

We used a national birth cohort of all children and young people (CYP) born in Scotland from 1997 onwards, developed from administrative health datasets linked to public health surveillance data on SARS-CoV-2 test results, originally constructed for the PICNIC study.^19^ CYP born in 1997 onwards were included. Birth registrations comprised the cohort spine, and CYP are linked over time and between databases using the Community Health Index (CHI) number, a unique personal identifier recorded at all interactions with the Scottish National Health Service (NHS). Table 1 summarises the databases and variables used in this study.

**Table 1.** Datasets and variables from the national Scottish birth cohort used in the study.

| **Dataset** | **Dataset details** | **Variables used** |
| --- | --- | --- |
| National Records for Scotland (NRS) birth registrations | Vital registration data on all children born in Scotland and their parents, collected via registry offices | Week and year of birth; Baby Sex; Socio-economic position (parents' occupation at birth). |
| Scottish Morbidity Record (SMR)-01 | Contains data on post-neonatal admissions and day cases to all NHS hospitals in Scotland | Admission and discharges dates; Primary and secondary diagnoses during admission; Type of hospital admission; Admission and discharge data from Intensive Care Unit (ICU). |
| SMR-02 (maternity records) | Contains data on all maternity admissions (including deliveries) in Scotland | Estimated gestational age; birth weight; number of older siblings (parity). |
| COVID-19 Tests | Contains data on all PCR- and antigen tests for SARS-Cov-2 with results and dates | Date of testing; Type of test; Result. |
| National Records for Scotland (NRS) death registrations | Vital registration data on children who died in Scotland | Date of death; Cause of death. |
| Scottish Birth Records (SBR) | Contains data on all children born in NHS hospitals, with data on neonatal admissions in and after April 2003 | Diagnoses recorded at or shortly after birth ; Primary and secondary diagnoses at birth admission. |
| CHI register | Contains data on migration in/out Scotland | Migration outside Scotland. |
| Child Health Surveillance Programme-School | Contains data school health visits | Height and weight at age 5 |

### Study population and follow-up

We included CYP born in Scotland from 1^st^ April 1997 to 31st December 2020. Children born at less than 24 completed weeks’ gestation or with a birthweight <500 grammes were excluded to ensure stillbirths were not inadvertently included.^20^ CYP whose mothers were not resident in Scotland at the time of delivery, and CYP who migrated out of Scotland before 1^st^ February 2020 were also excluded. For analyses of SARS-CoV-2 tests and positive test results (from now on referred to PCR-confirmed infections), CYP were followed from birth or the 1^st^ August 2020 (whichever occurred last), until death, migration from Scotland, or their 23^rd^ birthday, whichever occurred first. 1^st^ August 2020 was chosen as the follow-up start date for analyses of tests and PCR-confirmed infections since this is when testing for SARS-CoV-2 became commonly available in the community (rather than solely in hospitals) for children of all ages.^21^ For calculation and analyses of rates of COVID-19-related admissions, we used the follow-up start date as the 1^st^ February 2020. This allowed us to include all COVID-19-related hospital admissions since PCR testing for SARS-CoV-2 was available for suspected cases in hospital from January 2020.

### Outcomes

Our primary outcomes were rates of a) SARS-CoV-2 PCR tests (positive or negative); b) PCR-confirmed SARS-CoV-2 infections; c) COVID19-related hospital admissions. We also considered as secondary outcomes d) PIMS-TS admissions and e) COVID19-related intensive care unit (ICU) stays.

For a), we included all SARS-CoV-2 PCR tests recorded between1^st^ August 2020 and 31^st^ December 2020 in the COVID19 Tests Dataset. The samples were collected in hospitals, primary care, via national testing centres, or self-collection via home test kits. We did not include antigen (lateral flow device) test results, as they were not comprehensively represented in the COVID19 Tests dataset during 2020 (only 5% of test results in the cohort during the study period were from lateral flow devices). We defined as duplicate tests multiple tests taken on the same day, in the same CYP, with the same result, irrespective of whether they were taken at different locations. All duplicate tests, whether positive or negative, were excluded when calculating testing rates. For b), a PCR-confirmed SARS-CoV-2 infection was defined as the first record of a positive SARS-CoV-2 PCR test result (the index positive test) recorded in the COVID19 Tests dataset between1^st^ August 2020 and 31^st^ December 2020. Public Health Scotland recommends excluding all repeat positive tests within 90 days of the index positive sample date, and less than 5 CYP had multiple positive results beyond this time period. Therefore, only the first positive SARS-CoV-2 PCR test result for each child was included when calculating rates of PCR-confirmed SARS-CoV-2 infections, and identifying COVID-19-related hospital admissions.

We included all COVID-19-related hospital admissions (outcome c) between 1^st^ February and 31^st^ December 2020. To define COVID-19 related hospital admissions, we first linked episodes in the hospital admission dataset (Scottish Morbidity Record-01; Table 1) into admissions by assuming that episodes where the difference between the admission date and previous discharge date was ≤1 day^22^ indicated the same admission. Second, we identified COVID-19 related admissions where: (i) an individual had tested positive for SARS-CoV-2 up to 14 days prior to hospital admission, on the day of admission, or in between the hospital admission and discharge date, and/or (ii) an International Classification of Diseases-version 10 (ICD-10) diagnostic code for COVID-19 (U07.1 – U07.2) had been recorded during an admission as a primary or secondary diagnosis.

Since the ICD-10 code for PIMS-TS (U07.5) was introduced at the end of the follow-up period, we used other ICD-10 codes indicating systemic inflammatory response syndrome of infectious origin without organ failure (R65X), cardiogenic shock (R57X) or other specified systemic involvement of connective tissue (M35.8), suggestive of PIMS-TS recorded during an admission which had a positive SARS-CoV-2 PCR test within 28 day prior to the admission date.

A COVID-19-related intensive care unit (ICU) stay (outcome e) was defined where a child had an SMR-01 episode with ‘significant facility’ recorded with a positive SARS-CoV-2 PCR test to 21 days prior to the start of, or during, the ICU stay. ICU episodes where the difference between the ICU admission date and previous ICU discharge date was ≤1 day were assumed to indicate the same ICU stay.

### Risk factors

We examined four risk key factors for outcomes a-c): age group, sex, family socio-economic position (SEP) and history of chronic conditions. Age group was defined on 1^st^ February 2020 as the following five categories: <1 year (this also includes children born during 2020), 1-4 years, 5-11 years, 12-17 years and 18-22 years. We chose these age groups to reflect likely mixing patterns based on age (i.e. prior to formal childcare, nursery/preschool, primary school, secondary school, and higher/further education or work). Sex was recorded on the birth record. Family SEP was defined using parents’ (father’s, or mother’s if the birth was not jointly registered) occupation recorded on birth registration. This was coded using the UK National Statistics Socio-economic Classification (NS-SEC).^23^ We collapsed the NS-SEC classes into three groups: high SEP (managerial and professional occupations), middle SEP (intermediate occupations) and low SEP (routine and manual occupations). We identified history of chronic conditions by examining ICD-10 diagnostic codes recorded in SMR-01 in the previous five years. For children aged less than five years old at the start February 2020 or born during 2020, we used all available SMR-01 data and any diagnoses recorded on Scottish Birth Records. The code list for identifying chronic conditions was developed by Hardelid et al;^24^ and classifies chronic conditions into 8 types depending on body system: developmental/mental health, blood/cancer, chronic infections, respiratory, metabolic/gastrointestinal/endocrine/genitourinary, musculoskeletal/skin neurological/sensory, and cardiac conditions. We grouped the history of chronic conditions as none, one type of condition, and more than type of chronic condition.

We further explored whether gestational age and the number of older siblings affected outcomes b) and c) in children aged <5 years, and BMI in CYP aged 5-17, after adjusting for the four key risk factors. Gestational age was coded into a binary variable: preterm (<37 weeks) and term/late term (≥37 weeks). Number of older siblings (indicated by parity recorded on maternity records) was grouped into a three-category variable: no older siblings, one older sibling and two or more older siblings. BMI was derived from the Child Health Surveillance Programme - School dataset, which includes height and weight measurements for children starting their first (reception) year at primary school (at age 5 years), categorised using the British 1990 growth reference standards^25^ as underweight (<5^th^ percentile), healthy weight (5^th^ to <85^th^ percentile) and overweight/obese (≥85^th^ percentile).

### Statistical analyses

We calculated rates of outcomes a) and b) per 1,000 CYP years and outcome c) per 100,000 CYP-years with 95% confidence intervals, according to each of the risk factors. We estimated the median length of stay with interquartile ranges (IQRs) for COVID-19 related hospital admissions.

We examined the association between risk factors and testing rates using Poisson regression models with robust standard errors to account for multiple tests per child. To examine the association between risk factors and PCR-confirmed SARS-CoV-2 infection, and COVID19-related admission risk we used Cox proportional hazards regression models. Where a child had multiple COVID-19-related admissions, only the first was included in the Cox proportional hazards models. For each primary outcome, we first fitted an overall model including all ages. We included age group, sex, SEP and history of chronic conditions a priori as risk factors in the main model. We tested for interaction with age group and each of the other main risk factors using the Wald test. Two-sided *p*-values <0.05 were considered statistically significant.

Based on these results, we then fitted models for each primary outcome stratified by age group if a statistically significant interaction with age was identified for any of the other variables or if we identified non-proportional hazards. In further analyses for ages <5 years old we included parity and gestational age as additional risk factors in the models; and for ages 5-17 years we included BMI category. We tested the proportional hazards assumption of the Cox model by inspecting plots of Schoenfeld residuals^26^ and survival curves according to each main risk factors.

As there was only a small number of events for our secondary outcomes, we did not carry out detailed statistical analyses. We report the number of cases, median length of stay and age (with IQRs) for these outcomes.

All analyses were based on complete cases, as only a small number of CYP and missing values for any of the main variables. All statistical analyses were performed using Stata 16.0.

### Sensitivity analyses

We repeated the analyses for outcome c) using a more specific definition of a COVID-19-related admission restricted to emergency admissions with U07.1 or U07.2^27^ as the primary diagnosis.

## Results

This study included 1,226,855 CYP (Figure 1). Baseline characteristics of the participants are presented in Table 2. The median age on February 2020 was 11 years (IQR 5-17), and 5.4% of the cohort (66,974/1,226,855 CYP) had at least one chronic condition recorded in their hospital or birth record in the previous five years.

**Figure 1.**
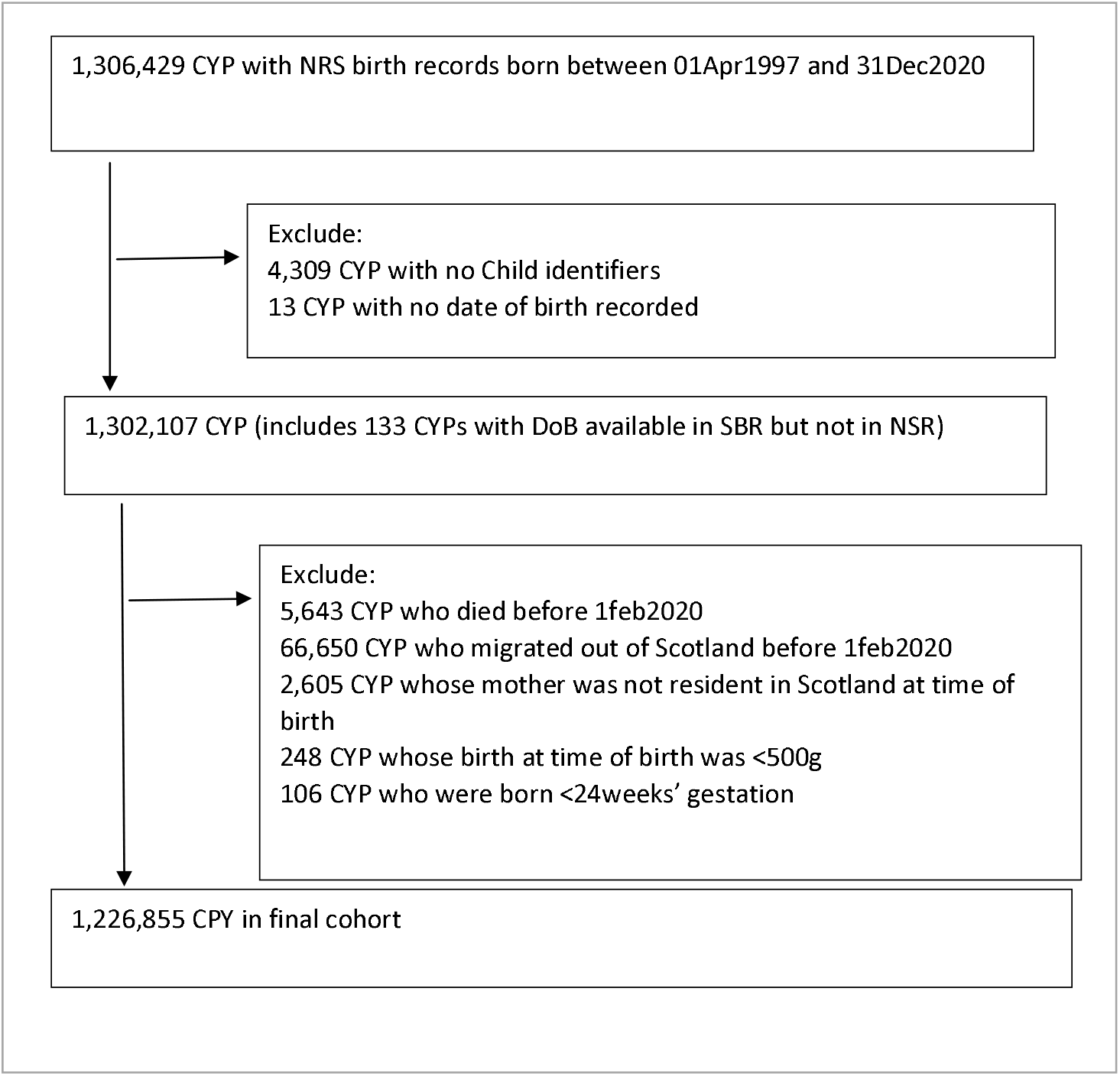
Flow chart describing creation of the final cohort

**Table 2.**
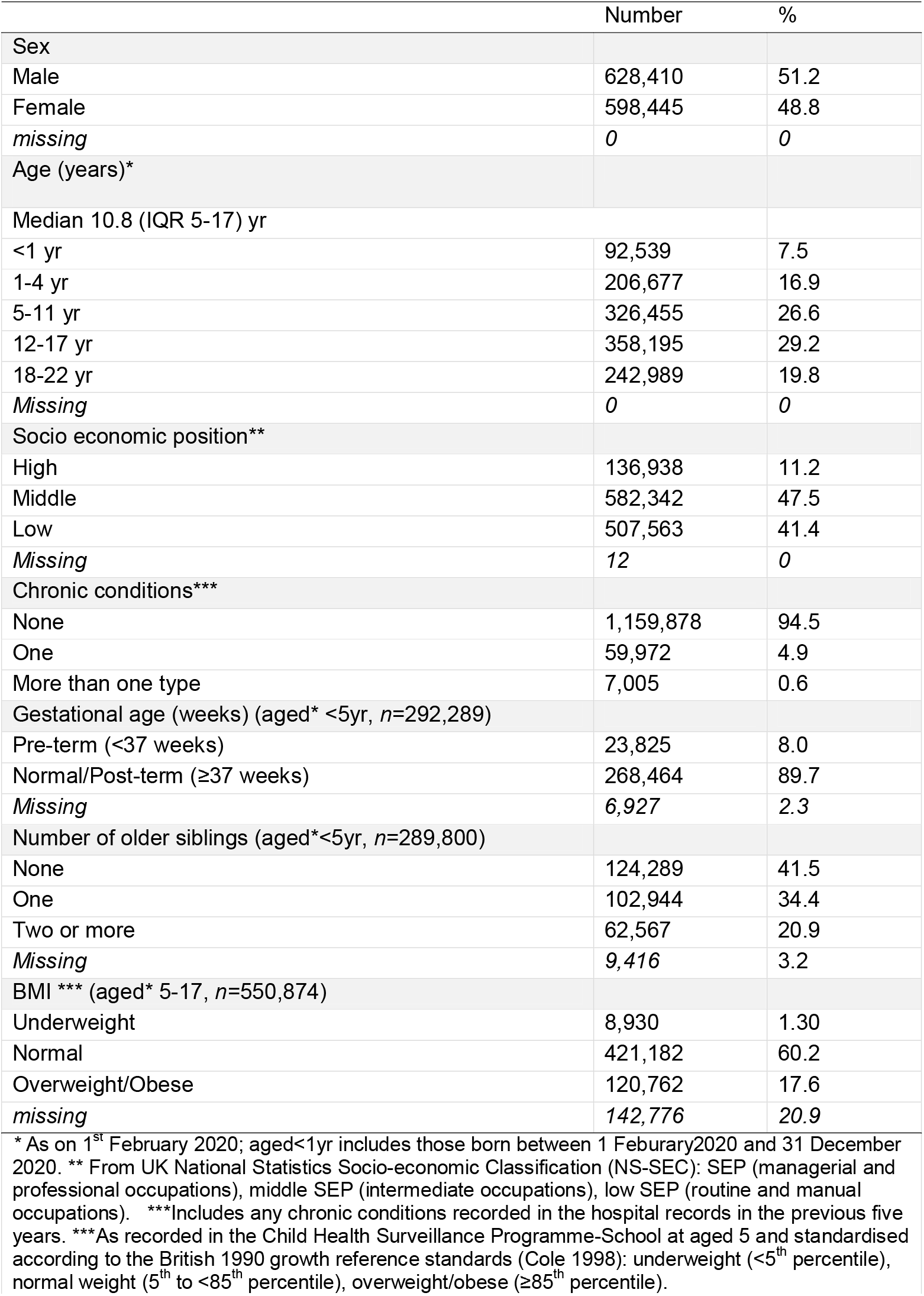
Cohort baseline characteristics (*n*=1,226,855)

|  | Number | % |
| --- | --- | --- |
| Sex |  |  |
| Male | 628,410 | 51.2 |
| Female | 598,445 | 48.8 |
| *missing* | 0 | 0 |
| Age (years)* |  |  |
| Median 10.8 (IQR 5-17) yr |  |  |
| <1 yr | 92,539 | 7.5 |
| 1-4 yr | 206,677 | 16.9 |
| 5-11 yr | 326,455 | 26.6 |
| 12-17 yr | 358,195 | 29.2 |
| 18-22 yr | 242,989 | 19.8 |
| *Missing* | 0 | 0 |
| Socio economic position** |  |  |
| High | 136,938 | 11.2 |
| Middle | 582,342 | 47.5 |
| Low | 507,563 | 41.4 |
| *Missing* | 12 | 0 |
| Chronic conditions*** |  |  |
| None | 1,159,878 | 94.5 |
| One | 59,972 | 4.9 |
| More than one type | 7,005 | 0.6 |
| Gestational age (weeks) (aged* <5yr, n=292,289) |  |  |
| Pre-term (<37 weeks) | 23,825 | 8.0 |
| Normal/Post-term (≥37 weeks) | 268,464 | 89.7 |
| *Missing* | 6,927 | 2.3 |
| Number of older siblings (aged* <5yr, n=289,800) |  |  |
| None | 124,289 | 41.5 |
| One | 102,944 | 34.4 |
| Two or more | 62,567 | 20.9 |
| *Missing* | 9,416 | 3.2 |
| BMI *** (aged* 5-17, n=550,874) |  |  |
| Underweight | 8,930 | 1.30 |
| Normal | 421,182 | 60.2 |
| Overweight/Obese | 120,762 | 17.6 |
| *missing* | 142,776 | 20.9 |

\* As on 1<sup>st</sup> February 2020; aged<1yr includes those born between 1 February2020 and 31 December 2020. \*\* From UK National Statistics Socio-economic Classification (NS-SEC): SEP (managerial and professional occupations), middle SEP (intermediate occupations), low SEP (routine and manual occupations). \*\*\*Includes any chronic conditions recorded in the hospital records in the previous five years. \*\*\*As recorded in the Child Health Surveillance Programme-School at aged 5 and standardised according to the British 1990 growth reference standards (Cole 1998): underweight (<5<sup>th</sup> percentile), normal weight (5<sup>th</sup> to <85<sup>th</sup> percentile), overweight/obese (≥85<sup>th</sup> percentile).

### SARS-CoV-2 testing

Between 1^st^ August (week 31) to 31^st^ December 2020 (week 52) we identified 378,402 PCR tests linked to 256,741 CYP; hence 20.9% of CYP in the cohort had at least one test. Figure 2 shows the weekly number of PCR tests by age group. The majority of CYP had been tested only once (200,288; 78.0%); 40,188 (15.7%) had been tested twice and 16,265 (6.3%) more than twice.

**Figure 2.**
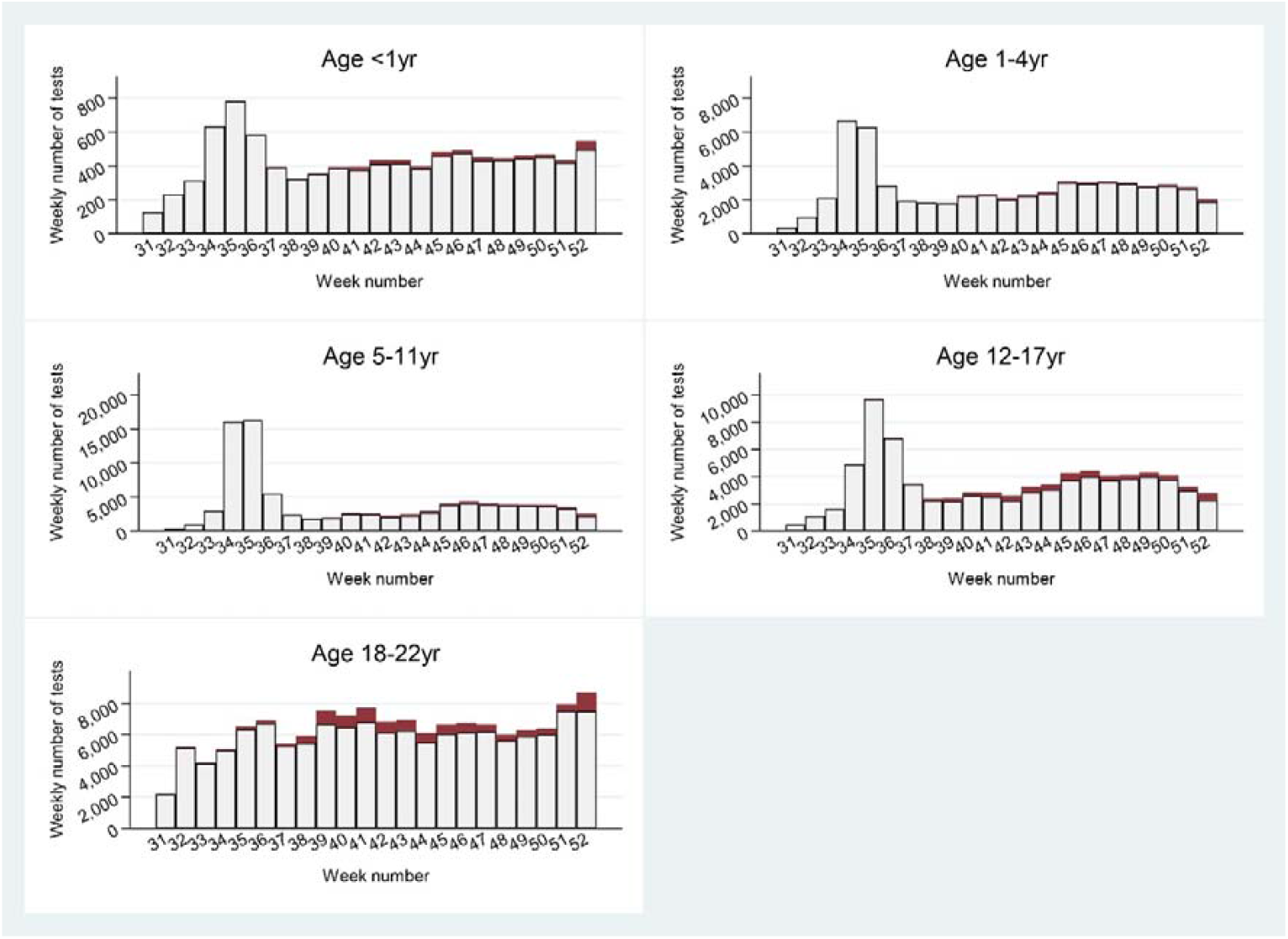
Weekly number of tests (white bars) and positive results (red bars) stratified by age group (weeks 31-52, 2020)

The crude testing rate was 770.8 (95%CI 768.4-773.3) per 1,000 CYP-years. Testing rates varied by age group and chronic conditions: children aged 1-4 years, young adults (age 18-22 years), and those with more than one chronic condition recorded had the highest testing rates (Appendix Table 1 & 2). Children aged <5 years born preterm were also more likely to be tested compared to children born at term (Appendix Table 3).

Models that mutually adjusted for all four main predictors suggested increasing age (and particularly an age of 18-22 years) and a history of chronic conditions were strongly associated with being tested (Table 3). As there was a statistically significant interaction term between age group and each of the other risk factors, we conducted age group-stratified analyses (Table 4). A history of chronic conditions was strongly associated with higher testing rates, with the size of the effect being the largest among infants

**Table 3.**
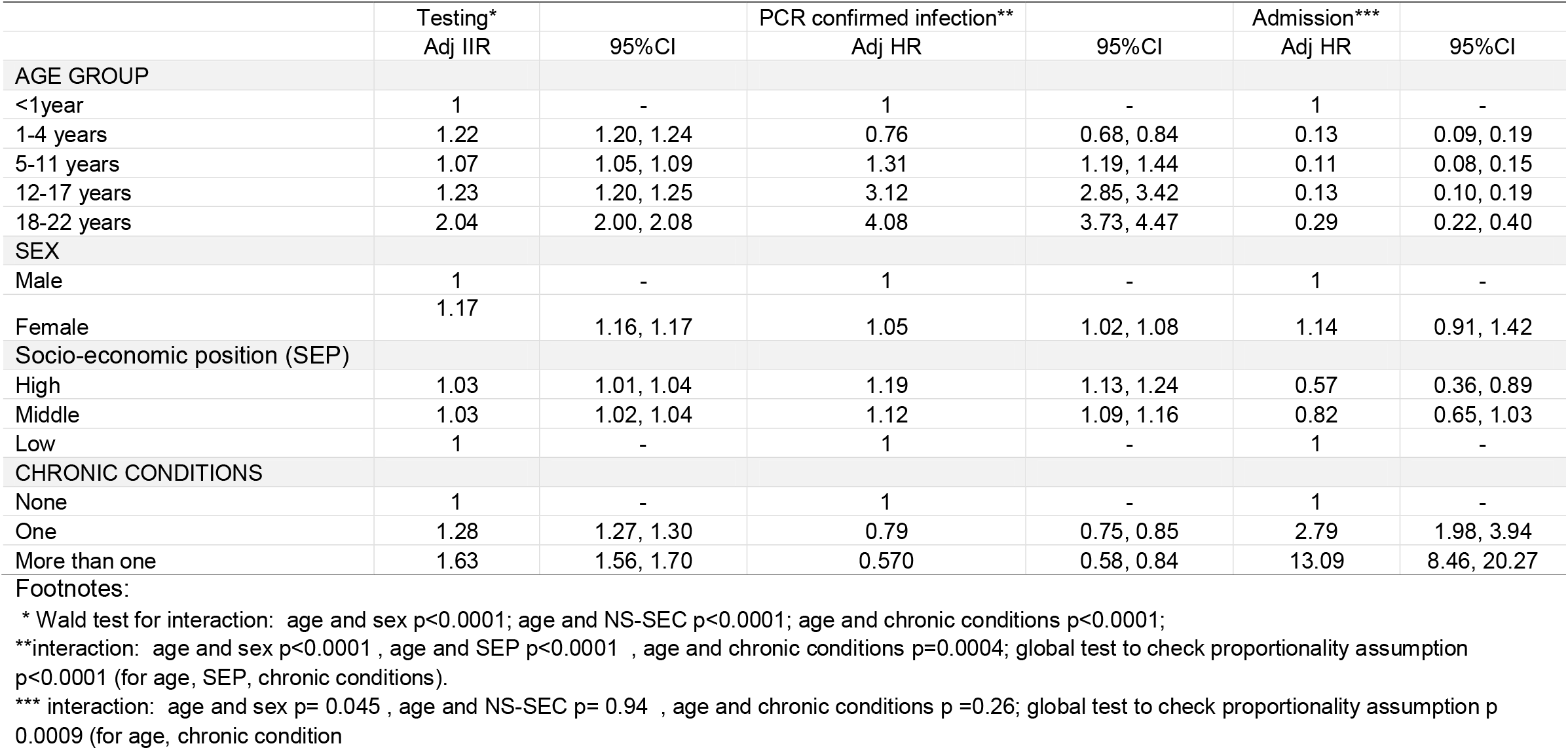
Results of models adjusted for age group, sex, socio-economic position and history of chronic conditions.

|  | Testing* |  | PCR confirmed infection** |  | Admission*** |  |
| --- | --- | --- | --- | --- | --- | --- |
|  | Adj IIR | 95%CI | Adj HR | 95%CI | Adj HR | 95%CI |
| **AGE GROUP** |  |  |  |  |  |  |
| <1year | 1 | - | 1 | - | 1 | - |
| 1-4 years | 1.22 | 1.20, 1.24 | 0.76 | 0.68, 0.84 | 0.13 | 0.09, 0.19 |
| 5-11 years | 1.07 | 1.05, 1.09 | 1.31 | 1.19, 1.44 | 0.11 | 0.08, 0.15 |
| 12-17 years | 1.23 | 1.20, 1.25 | 3.12 | 2.85, 3.42 | 0.13 | 0.10, 0.19 |
| 18-22 years | 2.04 | 2.00, 2.08 | 4.08 | 3.73, 4.47 | 0.29 | 0.22, 0.40 |
| **SEX** |  |  |  |  |  |  |
| Male | 1 | - | 1 | - | 1 | - |
| Female | 1.17 | 1.16, 1.17 | 1.05 | 1.02, 1.08 | 1.14 | 0.91, 1.42 |
| **Socio-economic position (SEP)** |  |  |  |  |  |  |
| High | 1.03 | 1.01, 1.04 | 1.19 | 1.13, 1.24 | 0.57 | 0.36, 0.89 |
| Middle | 1.03 | 1.02, 1.04 | 1.12 | 1.09, 1.16 | 0.82 | 0.65, 1.03 |
| Low | 1 | - | 1 | - | 1 | - |
| **CHRONIC CONDITIONS** |  |  |  |  |  |  |
| None | 1 | - | 1 | - | 1 | - |
| One | 1.28 | 1.27, 1.30 | 0.79 | 0.75, 0.85 | 2.79 | 1.98, 3.94 |
| More than one | 1.63 | 1.56, 1.70 | 0.570 | 0.58, 0.84 | 13.09 | 8.46, 20.27 |

\* Wald test for interaction: age and sex $p < 0.0001$ ; age and NS-SEC $p < 0.0001$ ; age and chronic conditions $p < 0.0001$ ;
\*\*interaction: age and sex $p < 0.0001$ , age and SEP $p < 0.0001$ , age and chronic conditions $p = 0.0004$ ; global test to check proportionality assumption $p < 0.0001$ (for age, SEP, chronic conditions).
\*\*\* interaction: age and sex $p = 0.045$ , age and NS-SEC $p = 0.94$ , age and chronic conditions $p = 0.26$ ; global test to check proportionality assumption $p$ $0.0009$ (for age, chronic condition)

**Table 4.** Incidence Risk Ratios of being <u>tested</u> by age group mutually adjusted for sex, socio-economic position and story of chronic conditions.

|  | Age<br><1 year |  | Age<br>1-4 years |  | Age<br>5-11 years |  | Age<br>12-17 years |  | Age<br>18-22years |  |
| --- | --- | --- | --- | --- | --- | --- | --- | --- | --- | --- |
|  |  |  | N |  | N |  | N |  | N |  |
| Number of CYP in model | 98333 |  | 216,048 |  | 397751 |  | 326249 |  | 317625 |  |
| N events | 9509 |  | 59176 |  | 92007 |  | 79771 |  | 137939 |  |
|  | Adj IIR | 95%CI | Adj IIR | 95%CI | Adj IIR | 95%CI | Adj IIR | 95%CI | Adj IIR | 95%CI |
| **SEX** |  |  |  |  |  |  |  |  |  |  |
| Male | 1 | - | 1 | - | 1 | - | 1 | - | 1 | - |
| Female | 0.88 | 0.85, 0.91 | 0.90 | 0.89, 0.92 | 0.90 | 0.89, 0.91 | 1.20 | 1.18, 1.22 | 1.61 | 1.58, 1.63 |
| **Socio economic position** |  |  |  |  |  |  |  |  |  |  |
| High | 1.59 | 1.52, 1.67 | 1.32 | 1.28, 1.35 | 0.96 | 0.94, 0.99 | 0.90 | 0.88, 0.93 | 0.93 | 0.91, 0.96 |
| Middle | 1.23 | 1.18, 1.27 | 1.11 | 1.09, 1.13 | 1.01 | 0.99, 1.02 | 0.99 | 0.97, 1.01 | 1.01 | 0.99, 1.03 |
| Low | 1 | - | 1 | - | 1 | - | 1 | - | 1 | - |
| **CHRONIC CONDITIONS** |  |  |  |  |  |  |  |  |  |  |
| None | 1 | - | 1 | - | 1 | - | 1 | - | 1 | - |
| One | 1.97 | 1.81, 2.15 | 1.30 | 1.25, 1.34 | 1.39 | 1.35, 1.43 | 1.37 | 1.32, 1.42 | 1.11 | 1.08, 1.13 |
| More than one | 3.53 | 3.20, 3.89 | 1.92 | 1.75, 2.10 | 2.13 | 1.95, 2.33 | 1.63 | 1.47, 1.80 | 1.12 | 1.05, 1.20 |

### PCR-confirmed infections

Among the 378,402 PCR tests identified in the cohort, 20,003 (5.3%) were positive and 7,275 (1.9%) were void. Excluding multiple positive tests per CYP, this corresponds to 19,005 PCR confirmed index infections in 7.4% (19,005/ 256,741) of the CYP who were tested between 1^st^ August 2020 and 31st December 2020.

The overall rate of PCR-confirmed infections was 179.4 (95% CI 176.9-182.0) per 1,000 CYP-years. Young adults (aged 18-22 years) had the highest rates of PCR-confirmed infections and those aged 1-4 years the lowest (Appendix Table 4 & 5). Infants had the highest PCR-confirmed infection rates in preschool children, otherwise infection rates were positively correlated with age. Among the infants who tested positive 20.3% (56/276) were aged <3months and 38.8% (107/276) aged <6months. Overall, CYP from higher SEP groups and those with no history of chronic conditions had a higher risk of PCR-confirmed infection (Table 3). As we identified statistically significant interaction terms between age group and each of the other main risk factors, we conducted stratified analyses by age group (Table 5). These results showed that among infants and preschool children, PCR-confirmed infection rates were lower among higher SEP groups, whereas among CYP aged 12 and above, children from higher SEP groups were more likely to be infected. Infants with chronic conditions had higher infection rates than those without; the opposite was true for CYP aged ≥12 years.

**Table 5.** Time to <u>PCR confirmed infection</u>: hazard ratios (HR) by age group mutually adjusted for sex, socio-economic position and history of chronic conditions.

|  | Age<br><1 years |  | Age<br>1-4 years |  | Age<br>5-11 years |  | Age<br>12-17 years |  | Age<br>18-22 years |  |
| --- | --- | --- | --- | --- | --- | --- | --- | --- | --- | --- |
| Number of CYP in model | 9661 |  | 49288 |  | 70245 |  | 76262 |  | 70212 |  |
| Number of PCR confirmed infections | 276 |  | 1114 |  | 1286 |  | 2706 |  | 4338 |  |
|  | AdjHR | 95%CI | AdjHR | 95%CI | AdjHR | 95%CI | AdjHR | 95%CI | AdjHR | 95%CI |
| **SEX** |  |  |  |  |  |  |  |  |  |  |
| Male | 1 | - | 1 | - | 1 | - | 1 | - | 1 | - |
| Female | 1.16 | 0.94, 1.42 | 1.08 | 0.97, 1.21 | 1.11 | 1.03, 1.19 | 1.20 | 1.14, 1.26 | 0.95 | 0.91, 0.99 |
| **SOCIO-ECONOMIC POSITION** |  |  |  |  |  |  |  |  |  |  |
| High | 0.70 | 0.49, 0.99 | 0.76 | 0.64, 0.92 | 0.86 | 0.75, 0.99 | 1.11 | 1.02, 1.20 | 1.52 | 1.43, 1.63 |
| Middle | 0.99 | 0.80, 1.24 | 0.99 | 0.88, 1.11 | 1.08 | 1.00, 1.17 | 1.13 | 1.07, 1.19 | 1.14 | 1.09, 1.19 |
| Low | 1 | - | 1 | - | 1 | - | 1 | - | 1 | - |
| **CHRONIC CONDITIONS** |  |  |  |  |  |  |  |  |  |  |
| None | 1 | - | 1 | - | 1 | - | 1 | - | 1 | - |
| One | 1.03 | 0.56, 1.88 | 0.80 | 0.63, 1.02 | 0.96 | 0.82, 1.14 | 0.86 | 0.77, 0.97 | 0.78 | 0.71, 0.84 |
| More than one | 2.04 | 1.08, 3.82 | 0.71 | 0.39, 1.29 | 0.86 | 0.52, 1.44 | 0.80 | 0.57, 1.13 | 0.42 | 0.41, 0.73 |

Additional analyses suggested that children aged <5 years with one older sibling had a reduced risk of a PCR confirmed infection compared to children with no older siblings (HR 0.63 (95%CI: 0.49-0.81) and 0.86 (95%CI: 0.76-0.98) for infants and 1-4 year olds, respectively, Appendix Table 6). Further, in children aged 12-17 years, being overweight/obese increased the risk of a PCR-confirmed infection compared to being of normal BMI (HR: 1.07 (95%CI: 1.00-1.15), Appendix Table 6).

### COVID19-related-hospital admissions

Between 1^st^ February 2020 and 31 December 2020, 55,940 CYP in the cohort were admitted to hospital at least once, accounting for 81,312 admissions. Of these admissions, 346 (0.6%) in 318 CYP were identified as COVID-19-related admissions. 25 CYP were admitted more than once. The median time to re-admission was 5 (IQR 2-12) days. The median length of stay was 2 days (IQR 1-4 days). There were 110 admissions between February and July (31.8%) and 236 (68.2%) between August and December (Figure 3).

**Figure 3.**
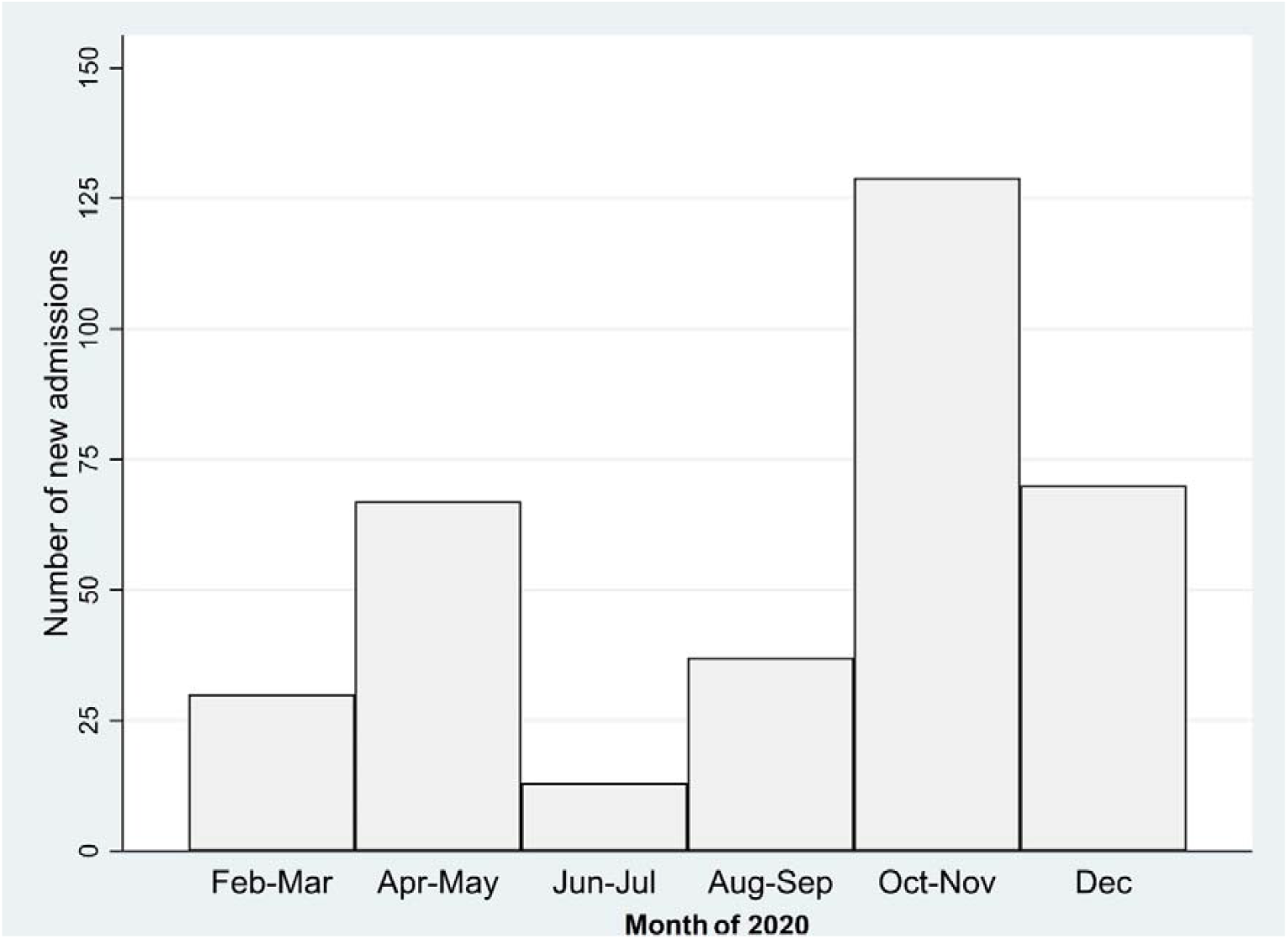
Monthly number of COVID-related hospital admissions (week 5-52, year 2020; 1^st^ February 2020 to 31^st^ December 2020)

Of the 346 COVID-19-related admissions 203 (58.7%) had a U07.1/U07.2 recorded as primary diagnosis and 258 (74.6%) were temporally associated with a SARS-CoV-2 positive test with or without a primary or secondary U07.1/U07.2 diagnosis (Figure 4).

**Figure 4.**
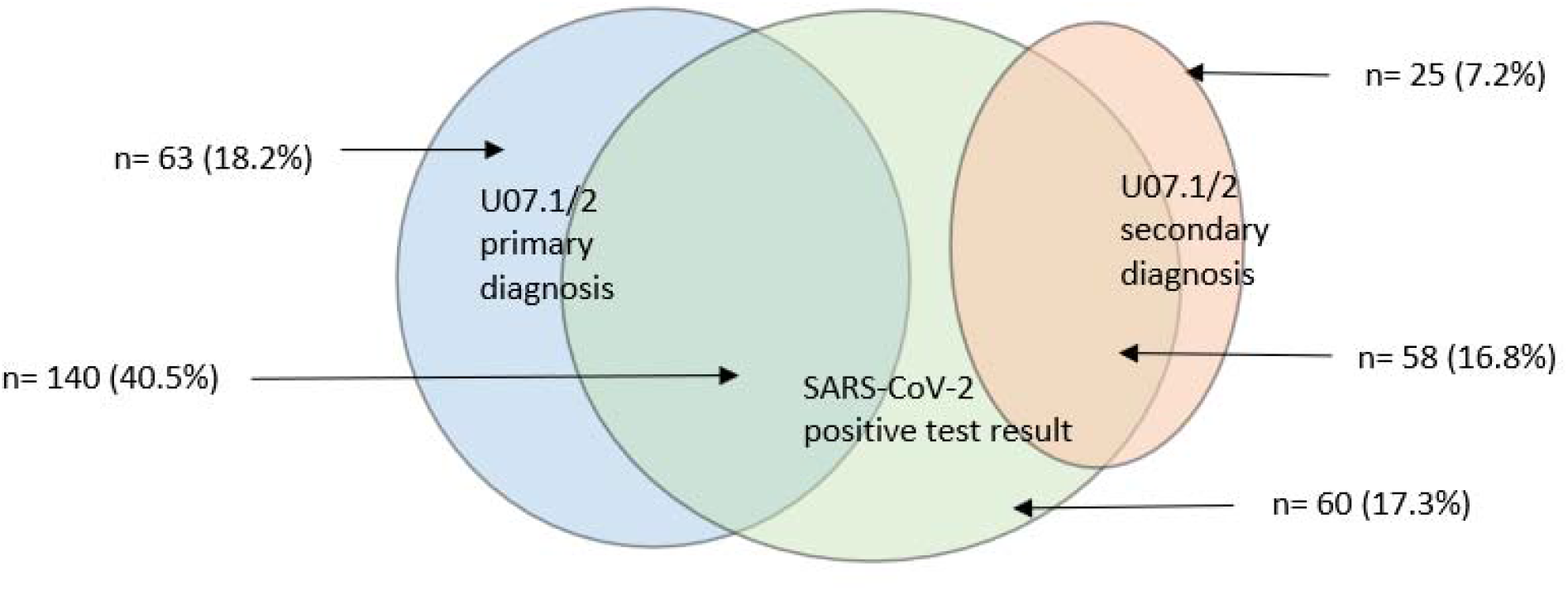
Number of COVID-related admissions temporally associated with PCR positive test up to 14 days before admission and by primary and secondary COVID19 diagnosis (U07.1/U07.2) Total admissions (*n)=* 346

The overall COVID-19 related admission rate was 29.4/100,000 (95%CI 26.3-32.8) CYP years. Infants had the highest COVID-19 related admission rate across the age groups: 120.6/100,000 (95%CI 92.2-157.9). CYP with more than one chronic condition type recorded had the highest admission rates across all age groups (Appendix table 7 & 8). Overall, 12.0% (*n*=38) of the 318 CYP admitted had at least one type of chronic condition, and 6.9% (*n*=22) had multiple types of chronic conditions recorded; thus 88% of admitted CYP did not have an underlying chronic condition.

Of the CYP with chronic conditions who had a COVID-19 related admission, neurological/sensory conditions were the common condition type recorded among children aged<12 years, whereas among those aged 12-22 years the most common conditions were developmental/mental health conditions followed by metabolic/gastrointestinal/endocrine/genitourinary conditions.

Across all ages, presence of one or more chronic condition increased the risk of COVID-19-related admissions and CYP from lower SEP groups had the highest risk of COVID-19-related admission (Table 3). We identified statistically significant interaction terms between age group and each of the other main risk factors, and therefore conducted age-stratified analyses (Table 6). Having more than one chronic condition remained the strongest risk factor for COVID-19-related admission across all age groups. There was no statistically significant association between prematurity, number of older siblings, or BMI category and COVID-19 related hospital admission risk (Appendix Table 9).

**Table 6.** Time-to-COVID related <u>admissions</u>: hazard ratios (HR) by age group mutually adjusted for sex, socio-economic position and history of chronic conditions.

|  | Age<br><1 year |  | Age<br>1-4 years |  | Age<br>5-11 years |  | Age<br>12-17 years |  | Age<br>18-22 years |  |
| --- | --- | --- | --- | --- | --- | --- | --- | --- | --- | --- |
| Number of<br>CYP in model | 92,530 |  | 251,884 |  | 347,542 |  | 385,664 |  | 268,467 |  |
| N Admissions | 53 |  | 51 |  | 55 |  | 49 |  | 110 |  |
|  | AdjHR | 95%CI | AdjHR | 95%CI | AdjHR | 95%CI | AdjHR | 95%CI | AdjHR | 95%CI |
| **SEX** |  |  |  |  |  |  |  |  |  |  |
| Male | 1 | - | 1 | - | 1 | - | 1 | - | 1 | - |
| Female | 0.81 | 0.51, 1.29 | 1.40 | 0.83, 2.36 | 0.96 | 0.54, 1.70 | 1.09 | 0.69, 1.72 | 1.47 | 0.99, 2.17 |
| **Socio-economic position** |  |  |  |  |  |  |  |  |  |  |
| High | 0.47 | 0.18, 1.19 | 0.47 | 0.17, 1.34 | 0.90 | 0.34, 2.38 | 0.31 | 0.10, 1.01 | 0.62 | 0.26, 1.45 |
| Middle | 0.82 | 0.51, 1.32 | 0.68 | 0.40, 1.17 | 0.91 | 0.49, 1.66 | 1.02 | 0.64, 1.63 | 0.74 | 0.50, 1.09 |
| Low | 1 | - | 1 | - | 1 | - | 1 | - | 1 | - |
| **CHRONIC CONDITIONS** |  |  |  |  |  |  |  |  |  |  |
| None | 1 | - | 1 | - | 1 | - | 1 | - | 1 | - |
| One | 1.37 | 0.34, 5.60 | 1.43 | 0.52, 3.98 | 3.30 | 1.40, 7.80 | 3.16 | 1.51, 6.61 | 3.75 | 2.34, 5.99 |
| More than one | 17.92 | 7.21, 44.55 | 12.87 | 5.12, 32.40 | 12.33 | 2.97, 51.17 | 20.93 | 9.03, 48.53 | 8.16 | 3.56, 18.75 |

### ICU admissions and PIMS-TS cases

Thirteen (3.8%) of the 346 COVID-related admissions involved an ICU attendance, accounting for 1.2% of the 1,238 ICU admissions in CYP during the study period. Around half of these admissions were in CYP with a history of one or more types of chronic conditions. The median age of CYP admitted to ICU was 14 years (IQR 9-19 years) and the median length of stay at ICU was 6 days (IQR 2-7 days). Half of COVID-related ICU admissions were in boys.

We identified less than five admissions with a diagnosis suggestive of PIMS-TS and temporally associated with a positive PCR test (<28 days prior admission by definition), all in males with an age spanning from 9 to 14 years. The median length of stay at admission was 10 days (IQR 6-14).

### Sensitivity analyses

Among the 346 COVID related admissions, 199 (57.5%) were emergency admissions with a primary diagnosis of U07.1/U07.2. Using the more specific definition, the COVID-19 related hospital admission rates were 107.0 (95% CI 80.4-142.4), 13.4 (9.0-19.8), 8.0 (5.6 – 11.7), 9.3 (6.3-13.6) and 27.7 (21.6-35.5) per 100,000 CYP years in age groups <1, 1-4, 5-11, 12-17 and 18-22 years respectively (Appendix Table 10 &11). This is 1.8 to 2 times higher than the more inclusive definition for CYP aged ≥1 year. Refitting the Cox proportional hazard models by age using the specific definition confirmed that presence of one or more chronic conditions remained the only significant risk factor for hospital admission (Appendix Tables 12 & 13). The median length of stay remained 2 days (IQR 1-5).

## Discussion

Over one fifth of CYP in Scotland had at least one SARS-CoV-2 PCR test during 2020, and 1.5% had a PCR-confirmed infection. Testing rates increased with increasing age. PCR-confirmed infection rates were highest in 18–22-year-olds, followed by secondary school children and infants. CYP with chronic conditions were more likely to be tested, but CYP with chronic conditions of secondary school age or older were less likely to have a PCR confirmed infection. Whilst COVID-19 related hospital admissions were uncommon (less than 3 per 10,000 CYP admitted in 2020), infants and those with multiple types of chronic conditions recorded had the highest COVID-19 related-admission rates. Preschool children from lower SEP groups were more likely to have PCR-confirmed infection, and secondary school children and young adults from lower SEP groups less likely. Across all ages, SEP was significantly associated with COVID-related hospital admission. We identified no statistically significant associations between prematurity, nor BMI and COVID-related admission risk; however the number of children admitted was small.

The well-established Scottish data linkage infrastructure allowed us to include data for all CYP born in Scotland since 1997, thereby minimising selection bias and loss to follow-up. We relied on linkage between hospital admission and public health surveillance data to define COVID-19-related admissions, allowing us to examine the robustness of our definitions in the linked data, rather than only relying on time difference between SARS-CoV-2 positive test and hospital admission alone. Importantly, by using a national birth cohort for this study, we could examine variations in population-based rates of SARS-CoV-2 testing, PCR confirmed infections and COVID-19-related hospital admissions across the full CYP age range, rather than only examining risk factors for ICU admission and death in hospitalised children. Using a population-wide study sample also minimises the risk of collider bias^28^ which may risk validity in studies only including hospitalised patients.

Despite the use of a national birth cohort, we were unable to determine risk factors for rarer outcomes including COVID-19-related ICU admission and cases of PIMS-TS. Pooling results across multiple countries with data linkage capabilities allowing national CYP birth cohorts to be created, could provide reliable risk estimates for these more severe outcomes following SARS-CoV-2 infection in CYP.

Further, this study included data from the first year of the pandemic, when wildtype (until November 2020), followed by Alpha (dominant from December 2020) SARS-CoV-2 variants were circulating in Scotland. This study will need to be repeated to examine the impact of new circulating variants, including Delta and Omicron, and changing transmission dynamics as vaccination of adults appear to be concentrating virus circulation among younger age groups.^29^ Recommendations to vaccinate high risk children aged 12 years and older against SARS-CoV-2 with two doses of Pfizer-BioNTech (BNT162b2; COMIRNATY) COVID vaccine was introduced in July 2021 in the UK; prior to this only children aged 12 and older with severe neurodisability, and 16-17 year olds with underlying conditions were recommended vaccination.^30^ In August and September 2021, these recommendations changed to advise vaccination with one dose for all 16-17 olds (due to risk of myocarditis), and then all children aged 12 years and above respectively.^31 32^ The US Food and Drug Administration approved the Pfizer-BioNTech vaccine for use in 5-11 year olds in late October 2021. Vaccination of children is likely to change risks of hospital admission, particularly among children with multiple chronic conditions. Whilst the benefits of vaccines may extend to non-health domains (e.g. school absences), further studies will need to examine whether the COVID-19 vaccination programme has amended the admission risks reported in this study. However, the results reported here provide a baseline during the first pandemic year against which more recent data can be compared.

We based our classification of chronic conditions on coded information in SMR-01 and SBR records; this may mean we have missed some conditions that are primarily managed in primary or community care settings. Our classification of socio-economic position was based on parental occupation derived from birth certificates, however, particularly among older CYP this may not reflect current socio-economic circumstances.

We highlight that infants have the highest admission risk among CYP, despite higher testing rates in older children and young adults. A previous systematic review (preprint) has indicated that infants are also at highest risk of requiring PICU admission once in hospital with COVID-19 disease.^14^ However, admission rates in infancy related to SARS-CoV-2 (1/1000 child-years) is lower than admission rates associated with confirmed influenza (2/1000 child-years)^33^ or respiratory syncytial virus infections (22/1000-child years).^34^ Future research should examine how COVID-19 vaccination programmes for pregnant women and older children, and removal of non-pharmaceutical interventions to control population mixing, affect infant SARS-CoV-2 admission rates.

We demonstrated that a history of chronic conditions, and particularly living with multiple different types of chronic conditions, was the most prominent risk factor for COVID-19 related hospital admission rates among CYP, however the vast majority of admitted children had no chronic conditions recorded. Children with chronic conditions also experience higher hospital admission rates for other conditions and injuries.^35^ CYP with chronic conditions were more likely to be tested than those without, however in older age groups CYP with chronic conditions were less likely to be positive. This may reflect lower threshold for testing among high-risk groups.

Preschool children from lower SEP groups (indicated by parental occupation recorded on the birth certificate) had higher risks of PCR-confirmed infection than children from higher SEP groups. This may be because younger children are spending more time in the home with their parents, and their risk of infection is therefore more strongly associated with their parents’ occupation (and ability to work from home). In older CYP, we instead identified higher PCR-confirmed infection rates among higher SEP groups, despite lower testing rates. This may be due to CYP from lower SEP groups being less likely to attend post-16 education, including university. Large outbreaks occurred in universities in Scotland in the autumn of 2020, which led to a surge in case numbers in 18-22 year olds.^36^ Linkage between SARS-CoV-2 test results, hospital admission and education data are required to confirm whether exposure in education settings can explain these differences in infection risk.

Low SEP at birth was also a risk factor for COVID-19-related admission in CYP in the overall model (incorporating all ages). This confirms previous reports which have indicated higher risk of severe outcomes in hospitalised adult COVID patients from more deprived areas,^37^ and higher all-age hospital admission rates in areas with higher area deprivation scores.^38 39^ Although we did not identify SEP as a significant risk factor for admission in age group specific models, COVID-19 related admission rates in children are much lower than in adults. Therefore, systematic differences in admission rates by SEP among specific age groups of CYP are harder to detect, even when using national data.

Our results showing that COVID-19 related admission rates in CYP peak in infancy indicates that further research and efforts to prevent COVID-19 admissions in children should include a focus on this age group. Pregnant women in Scotland are recommended to receive two doses of Pfizer/BioNTech COVID-19 vaccine. ^40^ As for pertussis^41 42^ and influenza,^43 44^ maternal vaccination during pregnancy could protect young babies from SARS-CoV-2 infection, however no studies to date have examined this. Further, given that CYP with chronic conditions are more likely to be admitted to hospital admission with COVID-19 disease than other CYP, studies monitoring the effectiveness of COVID-19 vaccines against severe outcomes in these high-risk groups are required to determine whether vaccination reduces the risk of admission.

We identified a peak in COVID-19-related hospital admissions in infants, and presence of chronic conditions as the strongest risk factor for hospital admissions in CYP, although the majority of admitted children did not have chronic conditions recorded. Further studies are urgently needed to examine whether maternal vaccine during pregnancy prevents COVID-19 admissions in infants. These data also provide baseline risks of infection and hospital admission for risk-benefit assessments of childhood vaccination, particularly for preschool children.

## Data Availability

The data used for this research are not openly available. Those wanting to access the data can apply to the electronic Data Research and Innovation Service (eDRIS) at Public Health Scotland: https://www.isdscotland.org/Products-and-services/Edris/

## Ethics statement

This study was approved by the University of Edinburgh School of Geosciences Ethics Committee (reference number 2020-401) and the Public Benefit and Privacy Panel for Health and Social Care (reference 1819-0049).

## Funding

UKRI-Medical Research Council

## Acknowledgements

We are grateful to Professor Bianca De Stavola (UCL) for her advice on statistical modelling, and Diane Rennie from Public Health Scotland for her help with data access and disclosure checking. This work uses data provided by patients and collected by the NHS as part of their care and support

## Conflicts of interest

None

## Notes

### Competing Interest Statement

The authors have declared no competing interest.

### Clinical Protocols

https://bmjopen.bmj.com/content/11/5/e048038

### Funding Statement

This study was funded by UK Research & Innovation Medical Research Council, grant reference number MR/T016558/1

### Author Declarations

This study was approved by the Public Benefit and Privacy Panel for Health and Social Care, reference number 1819-0049, and the University of Edinburgh School of Geosciences Ethics Committee, reference number 401.

### Summary of Updates

Paragraph 6 of the discussion has been updated to remove duplication of information

