## Supplementary tables for "Risk of SARS-CoV-2 testing, PCR-confirmed infections and COVID-19--related hospital admissions in children and young people: birth cohort study"

**Appendix: additional tables**

[**Table 3 SM Incidence Risk Ratio of being tested by age group mutually adjusted for sex, socio-economic status, history of chronic conditions and parity and pre-term (age<5 years) and BMI (aged 5-17 years) =**	4](#_Toc89784152)

[**Table 6 SM Time-to- PCR confirmed infection: hazard ratios (HR) by age group mutually adjusted for sex, socio-economic status, history of chronic conditions and parity and pre-term (age<5 years) and BMI (age 5-17 years)**	7](#_Toc89784155)

[**Table 9 SM Time-to-COVID related admission: hazard ratios (HR) by age group mutually adjusted for sex, socio-economic status, history of chronic conditions and parity and pre-term (age<5 years) and BMI (aged 5-17 years)**	10](#_Toc89784158)

**Appendix Table 1 Rate testing by age group (age 0-4 years as in Main Table 2 ) per 1,000 CYP-years**

|  | **Age <1 year** | | | | **Age 1-4 years** | | | |
| --- | --- | --- | --- | --- | --- | --- | --- | --- |
|  | Events | Rate | 95%LCI | 95%UCI | Events | Rate | 95%LCI | 95%UCI |
| Overall | 9509 | 482 | 473 | 492 | 59176 | 702 | 696 | 708 |
| SEX |  |  |  |  |  |  |  |  |
| Male | 5256 | 519 | 506 | 534 | 32284 | 743 | 735 | 752 |
| Female | 4253 | 443 | 430 | 457 | 26892 | 658 | 650 | 666 |
| Socio-economic position |  |  |  |  |  |  |  |  |
| High | 1368 | 551 | 522 | 581 | 9417 | 937 | 918 | 956 |
| Middle | 4417 | 466 | 452 | 480 | 29119 | 729 | 720 | 737 |
| Low | 3724 | 481 | 466 | 497 | 20637 | 602 | 594 | 610 |
| CHRONIC CONDITIONS |  |  |  |  |  |  |  |  |
| None | 8657 | 448 | 439 | 458 | 54124 | 682 | 677 | 688 |
| One | 489 | 1450 | 1327 | 1584 | 4049 | 922 | 894 | 950 |
| More than one | 363 | 5157 | 4652 | 5715 | 1003 | 1682 | 1581 | 1790 |
| GESTATIONAL AGE |  |  |  |  |  |  |  |  |
| pre-term | 8099 | 457 | 448 | 468 | 52447 | 693 | 687 | 699 |
| Term/post-term | 1207 | 797 | 753 | 843 | 5566 | 818 | 796 | 839 |
| NUMBER OLDER SIBLINGS |  |  |  |  |  |  |  |  |
| None | 4030 | 479 | 464 | 494 | 26920 | 770 | 761 | 779 |
| One | 3182 | 489 | 472 | 506 | 20177 | 692 | 682 | 701 |
| More than one | 1996 | 487 | 466 | 509 | 10371 | 589 | 577 | 600 |
| BMI |  |  |  |  |  |  |  |  |
| underweight | - | - | - | - | - | - | - | - |
| normal | - | - | - | - | - | - | - | - |
| overweight/obese | - | - | - | - | - | - | - | - |

**Appendix Table 2 Rate of testing by age group (age 5-22 years) per 1,000 CYP-years**

|  | **Age 5-11 years** | | | | **Age 12-17 years** | | | | **Age 18-22 years** | | | |
| --- | --- | --- | --- | --- | --- | --- | --- | --- | --- | --- | --- | --- |
|  | Events | Rate | 95%LCI | 95%UCI | Events | Rate | 95%LCI | 95%UCI | Events | Rate | 95%LCI | 95%UCI |
| Overall | 92007 | 583 | 580 | 587 | 79771 | 623 | 619 | 627 | 137939 | 1364 | 1357 | 1371 |
| SEX |  |  |  |  |  |  |  |  |  |  |  |  |
| Male | 49757 | 622 | 616 | 628 | 45322 | 586 | 581 | 592 | 46290 | 895 | 887 | 903 |
| Female | 42250 | 558 | 553 | 564 | 46831 | 635 | 630 | 641 | 91649 | 1854 | 1842 | 1866 |
| Socio-economic position |  |  |  |  |  |  |  |  |  |  |  |  |
| High | 10528 | 591 | 579 | 603 | 10059 | 531 | 521 | 541 | 9961 | 1198 | 1175 | 1222 |
| Middle | 41274 | 593 | 587 | 600 | 39678 | 612 | 606 | 618 | 78068 | 1390 | 1380 | 1399 |
| Low | 40203 | 588 | 582 | 595 | 42416 | 631 | 625 | 637 | 49909 | 1362 | 1350 | 1374 |
| CHRONIC CONDITIONS |  |  |  |  |  |  |  |  |  |  |  |  |
| None | 73955 | 577 | 572 | 581 | 85817 | 595 | 591 | 599 | 124563 | 1334 | 1327 | 1342 |
| One | 4880 | 819 | 797 | 843 | 5437 | 884 | 860 | 907 | 11952 | 1724 | 1694 | 1755 |
| More than one | 790 | 1493 | 1392 | 1601 | 899 | 1296 | 1214 | 1384 | 1424 | 1657 | 1573 | 1745 |
| GESTATIONAL AGE |  |  |  |  |  |  |  |  |  |  |  |  |
| pre-term | - | - | - | - | - | - | - | - | - | - | - | - |
| term | - | - | - | - | - | - | - | - | - | - | - | - |
| post-term | - | - | - | - | - | - | - | - | - | - | - | - |
| NUMBER OLDER SIBLINGS |  |  |  |  |  |  |  |  |  |  |  |  |
| None | - | - | - | - | - | - | - | - | - | - | - | - |
| One | - | - | - | - | - | - | - | - | - | - | - | - |
| More than one | - | - | - | - | - | - | - | - | - | - | - | - |
| BMI |  |  |  |  |  |  |  |  |  |  |  |  |
| underweight | 1009 | 626 | 589 | 666 | 1250 | 644 | 610 | 681 | - | - | - | - |
| normal | 49281 | 567 | 562 | 572 | 46792 | 594 | 589 | 600 | - | - | - | - |
| overweight/obese | 15287 | 588 | 579 | 598 | 14415 | 639 | 628 | 649 | - | - | - | - |

**Appendix Table 3 Incidence Risk Ratio of being tested by age group mutually adjusted for sex, socio-economic status, history of chronic conditions and parity and pre-term (age<5 years) and BMI (aged 5-17 years)**

|  | **Age <1 year** | | | **Age 1-4 years** | | | **Age 5-11 years** | | | **Age 12-17 years** | | |
| --- | --- | --- | --- | --- | --- | --- | --- | --- | --- | --- | --- | --- |
| *N* CYP in model | 89202 | | | 200590 | | | 310670 | | | 231202 | | |
| *N* tests in model | 94799 | | | 213258 | | | 323020 | | | 247143 | | |
|  | IRR | 95%LCI | 95%UCI | IRR | 95%LCI | 95%UCI | IRR | 95%LCI | 95%UCI | IRR | 95%LCI | 95%UCI |
| SEX |  |  |  |  |  |  |  |  |  |  |  |  |
| Male | 1.00 | - | - | 1.00 | - | - | 1.00 | - | - | 1.00 | - | - |
| Female | 0.88 | 0.85 | 0.91 | 0.91 | 0.89 | 0.92 | 0.90 | 0.89 | 0.91 | 1.16 | 1.13 | 1.18 |
| SOCIO-ECONOMIC POSITION | |  |  |  |  |  |  |  |  |  |  |  |
| High | 1.57 | 1.50 | 1.65 | 1.31 | 1.27 | 1.34 | 0.99 | 0.96 | 1.01 | 0.90 | 0.88 | 0.93 |
| Middle | 1.21 | 1.17 | 1.26 | 1.10 | 1.08 | 1.12 | 1.01 | 1.00 | 1.03 | 0.99 | 0.97 | 1.02 |
| Low | 1.00 | - | - | 1.00 | - | - | 1.00 | - | - | 1.00 | - | - |
| CHRONIC CONDITIONS | |  |  |  |  |  |  |  |  |  |  |  |
| None | 1.00 | - | - | 1.00 | - | - | 1.00 | - | - | 1.00 | - | - |
| One | 1.89 | 1.73 | 2.06 | 1.28 | 1.24 | 1.33 | 1.35 | 1.30 | 1.39 | 1.37 | 1.32 | 1.43 |
| More than one | 3.19 | 2.86 | 3.57 | 1.82 | 1.65 | 2.01 | 1.83 | 1.62 | 2.06 | 1.47 | 1.30 | 1.66 |
| GESTATIONAL AGE |  |  |  |  |  |  |  |  |  |  |  |  |
| Pre-term | 1.19 | 1.13 | 1.25 | 1.11 | 1.08 | 1.15 | - | - | - | - | - | - |
| Term/post-term | 1.00 | - | - | 1.00 | - | - | - | - | - | - | - | - |
| NUMBER OLDER SIBLINGS | |  |  |  |  |  |  |  |  |  |  |  |
| None | 1.00 | - | - | 1.00 | - | - | - | - | - | - | - | - |
| One | 0.98 | 0.95 | 1.02 | 0.91 | 0.89 | 0.93 | - | - | - | - | - | - |
| More than one | 0.87 | 0.83 | 0.91 | 0.83 | 0.81 | 0.85 | - | - | - | - | - | - |
| BMI |  |  |  |  |  |  |  |  |  |  |  |  |
| underweight | - | - | - | - | - | - | 1.06 | 0.99 | 1.12 | 1.10 | 1.02 | 1.19 |
| normal | - | - | - | - | - | - | 1.00 | - | - | 1.00 | - | - |
| overweight/obese | - | - | - | - | - | - | 1.03 | 1.01 | 1.05 | 1.06 | 1.03 | 1.08 |

**Appendix Table 4 Rate of PCR confirmed infections by age group (age 0-4 years) per 1,000 CYP-years**

|  | **Age<1 year** | | | | **Age 1-4 years** | | | |
| --- | --- | --- | --- | --- | --- | --- | --- | --- |
|  | Events | Rate | 95%LCI | 95%UCI | Events | Rate | 95%LCI | 95%UCI |
|  | 223 | 80 | 70 | 91 | 1136 | 62 | 58 | 65 |
| SEX |  |  |  |  |  |  |  |  |
| Male | 121 | 79 | 66 | 94 | 595 | 60 | 55 | 65 |
| Female | 102 | 81 | 67 | 98 | 541 | 64 | 59 | 69 |
| SOCIO-ECONOMIC POSITION |  |  |  |  |  |  |  |  |
| High | 21 | 51 | 33 | 78 | 141 | 51 | 43 | 60 |
| Middle | 111 | 83 | 69 | 100 | 571 | 63 | 58 | 69 |
| Low | 91 | 88 | 71 | 108 | 424 | 64 | 58 | 70 |
| CHRONIC CONDITIONS |  |  |  |  |  |  |  |  |
| None | 209 | 78 | 68 | 90 | 1065 | 62 | 59 | 66 |
| One | 7 | 81 | 39 | 171 | 60 | 51 | 39 | 65 |
| More than one | 7 | 188 | 90 | 395 | 11 | 55 | 31 | 99 |
| GESTATIONAL AGE |  |  |  |  |  |  |  |  |
| pre-term | 192 | 78 | 68 | 90 | 1022 | 62 | 59 | 66 |
| term/post-term | 24 | 85 | 57 | 126 | 89 | 53 | 43 | 66 |
| NUMBER OLDER SIBLINGS |  |  |  |  |  |  |  |  |
| None | 109 | 92 | 76 | 110 | 541 | 65 | 60 | 71 |
| One | 58 | 60 | 46 | 77 | 358 | 57 | 51 | 63 |
| More than one | 48 | 86 | 65 | 114 | 201 | 61 | 53 | 70 |
| BMI |  |  |  |  |  |  |  |  |
| underweight | - | - | - | - | - | - | - | - |
| normal | - | - | - | - | - | - | - | - |
| overweight/obese | - | - | - | - | - | - | - | - |

**Appendix** **Table 5 Rate of PCR confirmed infections (age 5-22 years) per 1,000 CYP-years**

|  | **Age 5-11 years** | | | | **Age 12-17 years** | | | | **Age 18-22 years** | | | |
| --- | --- | --- | --- | --- | --- | --- | --- | --- | --- | --- | --- | --- |
|  | Events | Rate | 95%LCI | 95%UCI | Events | Rate | 95%LCI | 95%UCI | Events | Rate | 95%LCI | 95%UCI |
|  | 3039 | 96 | 93 | 100 | 4929 | 193 | 188 | 198 | 9687 | 350 | 344 | 358 |
| SEX |  |  |  |  |  |  |  |  |  |  |  |  |
| Male | 1545 | 91 | 87 | 96 | 2313 | 179 | 172 | 187 | 4487 | 365 | 355 | 376 |
| Female | 1494 | 102 | 97 | 108 | 2616 | 207 | 199 | 215 | 5200 | 338 | 329 | 348 |
| SOCIO-ECONOMIC POSITION |  |  |  |  |  |  |  |  |  |  |  |  |
| High | 292 | 81 | 73 | 91 | 503 | 175 | 161 | 191 | 1181 | 481 | 455 | 510 |
| Middle | 1446 | 102 | 97 | 108 | 2231 | 202 | 194 | 210 | 5510 | 353 | 344 | 363 |
| Low | 1301 | 94 | 89 | 100 | 2195 | 189 | 181 | 197 | 2996 | 312 | 301 | 324 |
| CHRONIC CONDITIONS |  |  |  |  |  |  |  |  |  |  |  |  |
| None | 2855 | 97 | 93 | 100 | 4695 | 196 | 190 | 201 | 9056 | 359 | 352 | 367 |
| One | 165 | 93 | 80 | 108 | 208 | 151 | 132 | 173 | 581 | 267 | 246 | 290 |
| More than one | 19 | 95 | 60 | 148 | 26 | 147 | 100 | 215 | 50 | 194 | 147 | 256 |
| GESTATIONAL AGE |  |  |  |  |  |  |  |  |  |  |  |  |
| pre-term | - | - | - | - | - | - | - | - | - | - | - | - |
| term | - | - | - | - | - | - | - | - | - | - | - | - |
| post-term | - | - | - | - | - | - | - | - | - | - | - | - |
| NUMBER OLDER SIBLINGS |  |  |  |  |  |  |  |  |  |  |  |  |
| None | - | - | - | - | - | - | - | - | - | - | - | - |
| One | - | - | - | - | - | - | - | - | - | - | - | - |
| More than one | - | - | - | - | - | - | - | - | - | - | - | - |
| BMI |  |  |  |  |  |  |  |  |  |  |  |  |
| underweight | 38 | 112 | 82 | 154 | 72 | 179 | 142 | 225 | - | - | - | - |
| normal | 1723 | 101 | 96 | 106 | 2791 | 182 | 175 | 189 | - | - | - | - |
| overweight/obese | 545 | 104 | 96 | 113 | 918 | 198 | 186 | 211 | - | - | - | - |

**Appendix Table 6 Time to PCR confirmed infection: hazard ratios (HR) by age group mutually adjusted for sex, socio-economic status, history of chronic conditions and parity and pre-term (age<5 years) and BMI (age 5-17 years)**

|  | **Age <1 years** | | | **Age 1-4 years** | | | **Age 5-11 years** | | | **Age 12-17 years** | | |
| --- | --- | --- | --- | --- | --- | --- | --- | --- | --- | --- | --- | --- |
| *N* CYP in model | 9396 | | | 47930 | | | 46753 | | | 61991 | | |
| *N* events in model |  | | |  | | |  | | |  | | |
|  | HR | 95%LCI | 95%UCI | HR | 95%LCI | 95%UCI | HR | 95%LCI | 95%UCI | HR | 95%LCI | 95%UCI |
| SEX |  |  |  |  |  |  |  |  |  |  |  |  |
| Male | 1.00 | - | - | 1.00 | - | - | 1.00 | - | - | 1.00 | - | - |
| Female | 1.15 | 0.93 | 1.42 | 1.09 | 0.98 | 1.22 | 1.09 | 1.00 | 1.19 | 1.21 | 1.14 | 1.28 |
| SOCIO-ECONOMIC Position |  |  |  |  |  |  |  |  |  |  |  |  |
| High | 0.70 | 0.48 | 1.00 | 0.75 | 0.62 | 0.91 | 0.88 | 0.75 | 1.03 | 1.05 | 0.96 | 1.16 |
| Middle | 0.98 | 0.78 | 1.23 | 0.97 | 0.86 | 1.10 | 1.09 | 0.99 | 1.19 | 1.12 | 1.05 | 1.19 |
| Low | 1.00 | - | - | 1.00 | - | - | 1.00 | - | - | 1.00 | - | - |
| CHRONIC CONDITIONS | |  |  |  |  |  |  |  |  |  |  |  |
| None | 1.00 | - | - | 1.00 | - | - | 1.00 | - | - | 1.00 | - | - |
| One | 1.01 | 0.54 | 1.91 | 0.84 | 0.66 | 1.07 | 0.96 | 0.78 | 1.16 | 0.86 | 0.75 | 0.99 |
| More than one | 2.38 | 1.24 | 4.57 | 0.72 | 0.39 | 1.35 | 1.25 | 0.69 | 2.26 | 0.79 | 0.50 | 1.24 |
| GESTATIONAL AGE | |  |  |  |  |  |  |  |  |  |  |  |
| pre-term | 0.75 | 0.51 | 1.11 | 0.84 | 0.68 | 1.03 | - | - | - | - | - | - |
| Term/post-erm | 1.00 | - | - | 1.00 | - | - | - | - | - | - | - | - |
| NUMBER OLDER SIBLINGS |  |  |  |  |  |  |  |  |  |  |  |  |
| None | 1.00 | - | - | 1.00 | - | - | - | - | - | - | - | - |
| One | 0.63 | 0.49 | 0.81 | 0.86 | 0.76 | 0.98 | - | - | - | - | - | - |
| More than one | 0.81 | 0.62 | 1.07 | 0.90 | 0.77 | 1.05 | - | - | - | - | - | - |
| BMI |  |  |  |  |  |  |  |  |  |  |  |  |
| underweight | - | - | - | - | - | - | 1.03 | 0.72 | 1.47 | 1.03 | 0.84 | 1.27 |
| normal | - | - | - | - | - | - | 1.00 | - | - | 1.00 | - | - |
| overweight/obese | - | - | - | - | - | - | 1.06 | 0.95 | 1.17 | 1.07 | 1.00 | 1.15 |

**Appendix Table 7 TABLE Rate of admissions (age 0-4 years) per 100,000 CYP-years**

|  | **Age<1 years** | | | | **Age 1-4 years** | | | |
| --- | --- | --- | --- | --- | --- | --- | --- | --- |
|  | Events | Rate | 95%LCI | 95%UCI | Events | Rate | 95%LCI | 95%UCI |
|  | 53 | 121 | 92 | 158 | 51 | 27 | 21 | 36 |
| SEX |  |  |  |  |  |  |  |  |
| Male | 30 | 133 | 93 | 190 | 22 | 23 | 15 | 35 |
| Female | 23 | 107 | 71 | 162 | 29 | 32 | 22 | 46 |
| Socio-economic position |  |  |  |  |  |  |  |  |
| High | * | 73 | 27 | 194 | * | 18 | 7 | 48 |
| Middle | * | 118 | 80 | 175 | * | 24 | 16 | 36 |
| Low | * | 139 | 93 | 207 | * | 34 | 23 | 50 |
| CHRONIC CONDITIONS |  |  |  |  |  |  |  |  |
| None | * | 115 | 87 | 152 | * | 24 | 18 | 33 |
| One | * | 0 | 0 | 0 | * | 30 | 10 | 94 |
| More than one | * | 2212 | 830 | 5892 | * | 383 | 159 | 920 |
| GESTATIONAL AGE |  |  |  |  |  |  |  |  |
| pre-term | 42 | 106 | 79 | 144 | 41 | 24 | 18 | 33 |
| term/post-term | 10 | 290 | 156 | 538 | 8 | 53 | 27 | 106 |
| NUMBER OLDER SIBLINGS |  |  |  |  |  |  |  |  |
| None | 24 | 129 | 86 | 192 | * | 26 | 17 | 40 |
| One | 16 | 109 | 67 | 178 | * | 25 | 15 | 40 |
| More than one | 11 | 120 | 67 | 217 | * | 31 | 17 | 54 |
| BMI |  |  |  |  |  |  |  |  |
| underweight | - | - | - | - | - | - | - | - |
| normal | - | - | - | - | - | - | - | - |
| overweight/obese | - | - | - | - | - | - | - | - |

*redacted due to small numbers in some groups

**Appendix** **Table 8 TABLE Rate of admissions (age 5-22 years) per 100,000 CYP-years**

|  | **Age 5-11 years** | | | | **Age 12-17 years** | | | | **Age 18-22 years** | | | |
| --- | --- | --- | --- | --- | --- | --- | --- | --- | --- | --- | --- | --- |
|  | Events | Rate | 95%LCI | 95%UCI | Events | Rate | 95%LCI | 95%UCI | Events | Rate | 95%LCI | 95%UCI |
|  | 55 | 16 | 12 | 21 | 49 | 18 | 13 | 23 | 110 | 49 | 41 | 67 |
| SEX |  |  |  |  |  |  |  |  |  |  |  |  |
| Male | 31 | 17 | 12 | 25 | 26 | 18 | 12 | 27 | 43 | 38 | 28 | 55 |
| Female | 24 | 14 | 9 | 21 | 23 | 17 | 11 | 25 | 67 | 61 | 48 | 91 |
| Socio-economic position |  |  |  |  |  |  |  |  |  |  |  |  |
| High | * | 12 | 5 | 29 | * | 9 | 3 | 27 | 6 | 34 | 15 | 75 |
| Middle | * | 15 | 10 | 22 | * | 18 | 12 | 28 | 55 | 43 | 33 | 69 |
| Low | * | 18 | 12 | 26 | * | 19 | 13 | 29 | 49 | 62 | 47 | 86 |
| CHRONIC CONDITIONS |  |  |  |  |  |  |  |  |  |  |  |  |
| None | * | 14 | 10 | 18 | * | 15 | 11 | 20 | 81 | 39 | 32 | 57 |
| One | * | 40 | 18 | 89 | * | 51 | 23 | 114 | 23 | 147 | 98 | 237 |
| More than one | * | 293 | 110 | 782 | * | 221 | 71 | 687 | 6 | 307 | 138 | 682 |
| GESTATIONAL AGE |  |  |  |  |  |  |  |  |  |  |  |  |
| pre-term | - | - | - | - | - | - | - | - | - | - | - | - |
| term | - | - | - | - | - | - | - | - | - | - | - | - |
| post-term | - | - | - | - | - | - | - | - | - | - | - | - |
| NUMBER OLDER SIBLINGS |  |  |  |  |  |  |  |  |  |  |  |  |
| None | - | - | - | - | - | - | - | - | - | - | - | - |
| One | - | - | - | - | - | - | - | - | - | - | - | - |
| More than one | - | - | - | - | - | - | - | - | - | - | - | - |
| BMI |  |  |  |  |  |  |  |  |  |  |  |  |
| underweight | * | 54 | 13 | 215 | * | 24 | 3 | 168 | - | - | - | - |
| normal | * | 13 | 9 | 19 | * | 12 | 8 | 18 | - | - | - | - |
| overweight/obese | * | 15 | 8 | 29 | * | 25 | 14 | 44 | - | - | - | - |

*redacted due to small numbers in some groups

**Appendix Table 9 Time-to-COVID related admission: hazard ratios (HR) by age group mutually adjusted for sex, socio-economic status, history of chronic conditions and parity and pre-term (age<5 years) and BMI (aged 5-17 years)**

|  | **Age <1 year** | | | **Age 1-4 years** | | | **Age 5-11 years** | | | **Age 12-17 years** | | |
| --- | --- | --- | --- | --- | --- | --- | --- | --- | --- | --- | --- | --- |
| *N* CYP in model | 89197 | | | 244438 | | | 235396 | | | 316309 | | |
| *N* events in model |  | | |  | | |  | | |  | | |
|  | HR | 95%LCI | 95%UCI | HR | 95%LCI | 95%UCI | HR | 95%LCI | 95%UCI | HR | 95%LCI | 95%UCI |
| SEX |  |  |  |  |  |  |  |  |  |  |  |  |
| Male | 1.00 | - | - | 1.00 | - | - | 1.00 | - | - | 1.00 | - | - |
| Female | 0.80 | 0.49 | 1.29 | 1.48 | 0.86 | 2.54 | 0.61 | 0.29 | 1.29 | 0.90 | 0.52 | 1.57 |
| SOCIO-ECONOMIC Position |  |  |  |  |  |  |  |  |  |  |  |  |
| High | 0.39 | 0.14 | 1.11 | 0.49 | 0.17 | 1.40 | 1.60 | 0.50 | 5.10 | 0.36 | 0.08 | 1.51 |
| Middle | 0.80 | 0.49 | 1.31 | 0.64 | 0.37 | 1.13 | 1.56 | 0.71 | 3.45 | 1.25 | 0.71 | 2.20 |
| Low | 1.00 | - | - | 1.00 | - | - | 1.00 | - | - | 1.00 | - | - |
| CHRONIC CONDITIONS | |  |  |  |  |  |  |  |  |  |  |  |
| None | 1.00 | - | - | - | - | - | 1.00 | - | - | 1.00 | - | - |
| One | 1.26 | 0.30 | 5.20 | 1.50 | 0.54 | 4.17 | 2.41 | 0.73 | 7.96 | 2.77 | 1.10 | 7.00 |
| More than one | 14.38 | 5.33 | 38.84 | 12.11 | 4.42 | 33.15 | - | - | - | 11.88 | 2.87 | 49.14 |
| GESTATIONAL AGE |  |  |  |  |  |  |  |  |  |  |  |  |
| pre-term | 1.74 | 0.88 | 3.44 | 1.28 | 0.57 | 2.91 | - | - | - | - | - | - |
| term | 1.00 | - | - | 1.00 | - | - | - | - | - | - | - | - |
| NUMBER OLDER SIBLINGS |  |  |  |  |  |  |  |  |  |  |  |  |
| None | 1.00 | - | - | 1.00 | - | - | - | - | - | - | - | - |
| One | 0.83 | 0.47 | 1.45 | 0.83 | 0.45 | 1.53 | - | - | - | - | - | - |
| More than one | 0.96 | 0.52 | 1.75 | 0.87 | 0.44 | 1.75 | - | - | - | - | - | - |
| BMI |  |  |  |  |  |  |  |  |  |  |  |  |
| underweight | - | - | - | - | - | - | 2.27 | 0.30 | 16.85 | 2.39 | 0.57 | 9.99 |
| normal | - | - | - | - | - | - | 1.00 | - | - | 1.00 | - | - |
| overweight/obese | - | - | - | - | - | - | 1.08 | 0.46 | 2.53 | 1.51 | 0.82 | 2.78 |

**Appendix Table 10 TABLE Rate of ‘specific’ admissions (age 0-4 years) per 100,000 CYP-years**

|  | **Age<1 years** | | | | **Age 1-4 years** | | | |
| --- | --- | --- | --- | --- | --- | --- | --- | --- |
|  | Events | Rate | 95%LCI | 95%UCI | Events | Rate | 95%LCI | 95%UCI |
|  | 47 | 107.0 | 80.4 | 142.4 | 25 | 13.4 | 9.0 | 19.8 |
| SEX |  |  |  |  |  |  |  |  |
| Male | 26 | 115.4 | 78.6 | 169.5 | 10 | 10.4 | 5.6 | 19.3 |
| Female | 21 | 98.1 | 64.0 | 150.5 | 15 | 16.6 | 10.0 | 27.5 |
| Socio-economic position |  |  |  |  |  |  |  |  |
| High | * | 72.8 | 27.3 | 193.9 | * | 9.0 | 2.3 | 36.0 |
| Middle | * | 104.0 | 68.5 | 157.9 | * | 12.5 | 6.9 | 22.5 |
| Low | * | 121.5 | 79.2 | 186.3 | * | 15.7 | 8.9 | 27.7 |
| CHRONIC CONDITIONS |  |  |  |  |  |  |  |  |
| None | * | 105.2 | 78.6 | 140.9 | * | 13.1 | 8.7 | 19.7 |
| One | * | 0.0 | - | - | * | 20.2 | 5.0 | 80.7 |
| More than one | * | 1105.8 | 276.6 | 4421.4 | * | 0.0 | - | - |
| GESTATIONAL AGE |  |  |  |  |  |  |  |  |
| pre-term | 38 | 96.2 | 70.0 | 132.2 | * | 13.1 | 8.6 | 19.9 |
| term/post-term | 8 | 231.7 | 115.9 | 463.3 | * | 20.0 | 6.4 | 61.9 |
| NUMBER OLDER SIBLINGS |  |  |  |  |  |  |  |  |
| None | 21 | 112.6 | 73.4 | 172.6 | * | 14.2 | 7.9 | 25.7 |
| One | 15 | 102.2 | 61.6 | 169.5 | * | 13.9 | 7.2 | 26.7 |
| More than one | 9 | 98.4 | 51.2 | 189.1 | * | 12.8 | 5.3 | 30.7 |
| BMI |  |  |  |  |  |  |  |  |
| underweight | - | - | - | - | - | - | - | - |
| normal | - | - | - | - | - | - | - | - |
| overweight/obese | - | - | - | - | - | - | - | - |

*redacted due to small numbers in some groups

**Appendix** **Table 11 Rate of ‘specific’ admissions (age 5-22 years) per 100,000 CYP-years**

|  | **Age 5-11 years** | | | | **Age 12-17 years** | | | | **Age 18-22 years** | | | |
| --- | --- | --- | --- | --- | --- | --- | --- | --- | --- | --- | --- | --- |
|  | Events | Rate | 95%LCI | 95%UCI | Events | Rate | 95%LCI | 95%UCI | Events | Rate | 95%LCI | 95%UCI |
|  | 28 | 8.0 | 5.6 | 11.7 | 26 | 9.3 | 6.3 | 13.6 | 62 | 27.7 | 21.6 | 35.5 |
| SEX |  |  |  |  |  |  |  |  |  |  |  |  |
| Male | 15 | 8.4 | 5.1 | 14.0 | 14 | 9.8 | 5.8 | 16.5 | 27 | 23.6 | 16.2 | 34.4 |
| Female | 13 | 7.7 | 4.4 | 13.2 | 12 | 8.8 | 5.0 | 15.5 | 35 | 31.9 | 22.9 | 44.4 |
| Socio-economic position |  |  |  |  |  |  |  |  |  |  |  |  |
| High | * | 4.9 | 1.2 | 19.6 | * | 2.9 | 0.4 | 20.3 | * | 16.9 | 5.4 | 52.3 |
| Middle | * | 8.4 | 4.9 | 14.5 | * | 10.0 | 5.7 | 17.6 | * | 22.1 | 15.2 | 32.0 |
| Low | * | 8.5 | 5.0 | 14.7 | * | 10.4 | 6.0 | 17.9 | * | 39.0 | 27.4 | 55.5 |
| CHRONIC CONDITIONS |  |  |  |  |  |  |  |  |  |  |  |  |
| None | * | 7.2 | 4.9 | 10.8 | * | 7.9 | 5.1 | 12.1 | * | 21.3 | 15.8 | 28.6 |
| One | * | 6.7 | 0.9 | 47.5 | * | 25.6 | 8.2 | 79.2 | * | 89.7 | 53.1 | 151.4 |
| More than one | * | 220.1 | 71.0 | 682.4 | * | 147.7 | 36.9 | 590.4 | * | 204.4 | 76.7 | 544.6 |
| GESTATIONAL AGE |  |  |  |  |  |  |  |  |  |  |  |  |
| pre-term | - | - | - | - | - | - | - | - | - | - | - | - |
| term | - | - | - | - | - | - | - | - | - | - | - | - |
| post-term | - | - | - | - | - | - | - | - | - | - | - | - |
| NUMBER OLDER SIBLINGS |  |  |  |  |  |  |  |  |  |  |  |  |
| None | - | - | - | - | - | - | - | - | - | - | - | - |
| One | - | - | - | - | - | - | - | - | - | - | - | - |
| More than one | - | - | - | - | - | - | - | - | - | - | - | - |
| BMI |  |  |  |  |  |  |  |  |  |  |  |  |
| underweight | * | 53.9 | 13.5 | 215.4 | * | 0.0 |  |  | - | - | - | - |
| normal | * | 6.5 | 3.8 | 11.2 | * | 5.9 | 3.2 | 11.0 | - | - | - | - |
| overweight/obese | * | 10.0 | 4.5 | 22.3 | * | 12.4 | 5.6 | 27.7 | - | - | - | - |

*redacted due to small numbers in some groups

**Appendix Table 12 Time-to-COVID related admission (specific definition): hazard ratios (HR) by age group mutually adjusted for sex, socio-economic status and history of chronic conditions (age 0-4)**

|  | **Age <1 year** | | | **Age 1-4 years** | | |
| --- | --- | --- | --- | --- | --- | --- |
| *N* CYP in model | 92530 | | | 251884 | | |
| *N* events in model | 47 | | | 25 | | |
|  | HR | 95%LCI | 95%UCI | HR | 95%LCI | 95%UCI |
| SEX |  |  |  |  |  |  |
| Male | 1.00 | - | - | 1.00 | - | - |
| Female | 0.77 | 0.45 | 1.30 | 1.82 | 0.83 | 3.98 |
| SOCIO-ECONOMIC Position |  |  |  |  |  |  |
| High | 0.63 | 0.24 | 1.65 | 0.57 | 0.13 | 2.55 |
| Middle | 0.90 | 0.52 | 1.55 | 0.93 | 0.43 | 2.05 |
| Low | 1.00 | - | - | 1.00 | - | - |
| CHRONIC CONDITIONS | |  |  |  |  |  |
| None | 1.00 | - | - | 1.00 | - | - |
| One | 0.83 | 0.11 | 5.97 | 1.40 | 0.33 | 5.90 |
| More than one | 8.67 | 2.11 | 35.60 | -* | - | - |

*There were no children in this category for this age group

**Appendix Table 13 Time-to-COVID related admission (specific definition): hazard ratios (HR) by age group mutually adjusted for sex, socio-economic status and history of chronic conditions (age 5-22)**

|  | **Age 5-11 years** | | | **Age 12-17 years** | | | **Age 18-22 years** | | |
| --- | --- | --- | --- | --- | --- | --- | --- | --- | --- |
| *N* CYP in model | 347542 | | | 385664 | | | 268467 | | |
| *N* events in model | 28 | | | 26 | | | 62 | | |
|  | HR | 95%LCI | 95%UCI | HR | 95%LCI | 95%UCI | HR | 95%LCI | 95%UCI |
| SEX |  |  |  |  |  |  |  |  |  |
| Male | 1.00 | - | - | 1.00 | - | - | 1.00 | - | - |
| Female | 1.08 | 0.49 | 2.41 | 1.01 | 0.53 | 1.92 | 1.29 | 0.77 | 2.14 |
| SOCIO-ECONOMIC Position |  |  |  |  |  |  |  |  |  |
| High | 0.79 | 0.17 | 3.62 | 0.20 | 0.03 | 1.48 | 0.47 | 0.14 | 1.54 |
| Middle | 1.21 | 0.52 | 2.80 | 0.97 | 0.50 | 1.87 | 0.56 | 0.33 | 0.93 |
| Low | 1.00 | - | - | 1.00 | - | - | 1.00 | - |  |
| CHRONIC CONDITIONS | |  |  |  |  |  |  |  |  |
| None | 1.00 | - | - | 1.00 | - | - | 1.00 | - |  |
| One | 1.03 | 0.14 | 7.67 | 2.32 | 0.71 | 7.61 | 4.08 | 2.22 | 7.48 |
| More than one | 23.49 | 5.49 | 100.57 | 27.15 | 9.55 | 77.21 | 9.64 | 3.46 | 26.88 |
